# Clinician and caregiver Perceptions of the FASD Diagnostic Journey: A Qualitative Study

**DOI:** 10.64898/2025.12.14.25342215

**Authors:** Jordan Hill-Rucker, Claire Coles, Julie Kable, Hannah Cooper

## Abstract

**Background:** Alcohol consumption and binge drinking during pregnancy have been rising in the US since 2006. Unfortunately, capacity to diagnose fetal alcohol spectrum disorder (FASD) remains suboptimal. Incorporating the lived experiences of affected populations improves service access and responsiveness, and enhances quality of care. This qualitative study thus explored barriers and facilitators in the FASD diagnostic journey from the perspectives of caregivers of children diagnosed with FASD and clinicians engaged in screening, evaluating, or diagnosing FASD or linking children to needed interventions.

**Methods:** Caregivers were biological, adoptive, foster, or other guardians of children recently diagnosed with FASD at a large teaching hospital system serving a major southeastern metropolitan area. Clinicians were providers at this same hospital system who screened, evaluated, or diagnosed children with FASD or linked them to needed interventions. Study staff conducted semi-structured qualitative interviews with participants that covered barriers and facilitators arising during this diagnostic journey. Thematic analysis methods were applied to identify patterns across transcripts.

**Results:** Eleven clinicians and 15 caregivers participated. Clinicians and caregivers reported that barriers to FASD evaluation and diagnosis included (1) cost; (2) wait times; and (3) prenatal alcohol exposure documentation, often shaped by stigma; no facilitators were identified. Facilitators to linkage to interventions were caregiver education on FASD symptoms, services to address these symptoms, and how to connect to these services. Barriers were absence of needed services, long travel distances to existing services, and cost.

**Conclusions:** In this sample, stigma, cost, and provider availability impeded the FASD diagnostic journey and linkage to care. Promising proposed federal legislation (i.e., the FASD Respect Act) targets these barriers, and thus holds potential to support evaluation, diagnoses, and linkage to care among children with FASD.

## Introduction

Even as rates of binge drinking and overall alcohol consumption rise during pregnancy in the United States (US), identification and diagnosis of fetal alcohol spectrum disorders (FASD) remains difficult. Population-level surveys indicate that alcohol consumption during pregnancy has been rising since 2011, with a two percentage point increase in binge drinking during pregnancy between 2011 and 2020, to 5.2%.(Gosdin 2022) Prenatal alcohol exposure (PAE) can cause FASD, which is characterized by an array of lifelong neurological, cognitive, behavioral, learning, and sometimes physical challenges.(Hoyme et al. 2016; Rangmar et al. 2015) Evidence-based early intervention activities and special education services in older children can remediate many of these challenges and support people living with FASD in their homes, schools and workplaces, and communities.(Popova et al. 2023)

Intervention is, however, predicated on diagnosis,(Wilhoit, Scott, and Simecka 2017) and evidence indicates that in the US and elsewhere most people living with FASD are not diagnosed in childhood or at all.(Bertrand, Floyd, and Weber 2005; May et al. 2018; Chasnoff, Wells, and King 2015; May et al. 2009) May et al (2018) evaluated all first graders in four US geographic regions for FASD. Only 1% of children identified with FASD through the study had been previously diagnosed. Under-diagnosis is pervasive even in populations with known risk for FASD: Chasnoff et al. (2015) assessed a sample of foster/adopted youth with behavioral issues for FASD and found that just 13.5% of those diagnosed by the project had been previously diagnosed. Absent diagnosis, children and their caregivers cannot access needed services that can transform behavioral, learning, occupational, and health trajectories across the lifecourse.(Gibson et al. 2025) Likewise, high rates of underdiagnosis in particular geographic areas (e.g., public health districts) preclude effective allocation of resources to reduce PAE and treat FASD in communities.

Regardless of health outcomes, incorporating affected populations’ lived experiences strengthens healthcare planning, improves service access and responsiveness, and enhances quality of care outcomes.(Bombard et al. 2018) A recent review of the FASD diagnostic journey identified 10 qualitative studies with people living with FASD and their caregivers.(Hayes et al. 2023) Barriers to diagnoses included primary care providers’ dismissal of caregiver concerns about FASD; limited availability of providers skilled in FASD evaluation and diagnosis; and long wait times. All but one study, however, was conducted outside the US.(Hayes et al. 2023) Themes may be markedly different in the US, where many families have suboptimal health insurance coverage and punitive laws governing alcohol use in pregnancy in many states may complicate querying and recording PAE.(US Census Bureau 2024; Yu et al. 2022; Oh et al. 2024)

The present qualitative analysis thus explores caregiver and clinician experiences of the FASD diagnostic journey and linkage to related services in one Georgia-based hospital system that includes the main provider of FASD evaluation and diagnoses in the state. The goal of this analysis is to inform efforts to improve this journey and ensure that children living with FASD and their caregivers connect rapidly with essential interventions.

## Methods

### Institutional Review Board Approval

The university’s Institutional Review Board approved study protocols.

### Participant Eligibility Criteria and Recruitment

This qualitative study sampled two groups of participants: (1) clinicians in a university-based healthcare system in Georgia metropolitan area who screened, evaluated, or diagnosed children for FASD, and/or linked children to FASD-related services, and (2) caregivers of children who recently were diagnosed with FASD or received FASD-related services within the healthcare system. This study was funded through a contract with the Centers for Disease Control and Prevention (CDC) and had to abide by federal guidelines on participant burden. As a result, we were limited to sampling clinicians who worked at a single university healthcare system in Georgia and caregivers of patients receiving services through this system.

Eligibility criteria for caregivers included: (1) being at least 18 years old; (2) being a biological parent, adoptive parent, foster parent, or other legal guardian of a child who had been diagnosed with FASD or received FASD-related services at the hospital system in the past 5 years; (3) living in the Georgia metropolitan area where the hospital system was located; and (4) being sufficiently fluent to complete the screener and consent in English.

Caregivers were recruited via flyers distributed by providers at the hospital’s Prenatal Exposure Clinic (PEC), and PEC patient recruitment rosters. PEC maintains a census of all caregivers of children who received services at the clinic and who consented to be contacted about future research opportunities. Clinic directors mailed and e-mailed a letter about the project to individuals on this roster who had sought services between 2018-2023 to alert them to the study and its purpose, and to indicate that study participation would not affect services. A study staff member then sorted the roster by the most recent PEC service date, and contacted caregivers in the order in which they appeared in the roster. As part of the consent process, all caregivers were informed that PEC would not know who chose to be screened or take part in the interview.

Clinicians were eligible to take part if they (1) screened, evaluated, or diagnosed children with FASD or linked children to FASD-related services; individuals had to have been in this role for at least the past 6 months; (2) had treated ≥3 patients with suspected or diagnosed FASD in the 12 months prior to being screened; (3) were currently employed by the University; and (4) were sufficiently fluent to complete the English-language screener and consent process.

We recruited clinicians via three methods. First, the PEC directors alerted PEC clinicians to the study and study staff contacted all PEC clinicians to describe it. Second, we used a sampling frame to randomly sample pediatricians at the university with relevant expertise. We created the sampling frame by scanning the directory of the university’s pediatric department and the roster of clinicians who referred patients to PEC for FASD evaluation. Third, we invited each enrolled clinician to refer other clinicians for us to screen. As part of the consent process, clinicians were informed that taking part in the screening or interview would not affect any aspect of their job.

### Assessment and Instruments

Interview guides: We gathered data via qualitative semi-structured one-on-one interviews. The clinician interview guide queried experiences of and barriers and facilitators to FASD screening, evaluation, diagnosis, and linkage to care (see Table 1 for definitions). The guide followed a modular format, with each module exploring one of the above processes (e.g., screening, evaluation), to allow the interviewer to explore only questions that were relevant to each participating clinician and skip others (e.g., a participant who said that they did not engage in screening or linkage to care would not be asked questions about those processes; see clinician Interview Guide in Appendix 1). The caregiver interview guide explored initial triggers to seeking help, the FASD diagnostic journey, diagnostic delays and challenges, information exchange, and services (see Caregiver Interview Guide in Appendix 1).

**Table 1.** Definitions of domains in the clinician interview guide

| Table 1. Definitions of domains in the clinician interview guide |  |
| --- | --- |
| Screening | A process designed to identify individuals who may have an FASD and thus require formal FASD evaluation. |
| Evaluation | A process that distinguishes people who have FASD from those who do not. This process might include gathering information on whether the biological parent has a history of alcohol consumption during pregnancy, tests for pediatric neurocognitive/behavioral functioning, identification of dysmorphic features such as facial dysmorphism, and investigation of prenatal and postnatal growth parameters. |
| Diagnosis | A clinical decision that a patient does (not) have FASD based on the evaluation. |
| Linkage to Care | Connecting patients diagnosed with FASD to needed interventions and/or coordinating care. |

### Interview Process

After obtaining informed consent, study staff members conducted interviews via HIPPA-compliant Zoom or in-person, based on participant preference. Interviews lasted 45-90 minutes. Clinicians received a $50 e-gift card and caregivers received a $75 e-gift card for participating. Interviews were audio-recorded and recordings were transcribed verbatim using NVivo’s audio-transcription service. Transcripts were checked for accuracy by the study team.

### Analysis

The study team used Braun and Clarke’s six-phase thematic analysis method to analyze the transcripts (i.e., 1) familiarizing oneself with the data; 2) generating initial codes; 3) searching for themes; 4) reviewing themes; 5) defining and naming themes; and 6) producing the report).(Braun and Clarke 2021) We developed a preliminary codebook by applying deductive and inductive methods to clinician transcripts and then iteratively revising the codebook as more transcripts were coded (See codebook in Appendix 2). We then expanded this codebook to encompass phenomena described in caregiver interviews. The first author applied the codes to each transcript using NVIVO 14. Memos were developed by the first and second authors to conceptualize themes (step 3) and iteratively review, define, and name them (steps 4 & 5). We intentionally explored variation in the clinician sample by volume of patients seen with suspected or confirmed FASD and also between caregivers and clinicians. Findings are identical across groups unless specified.

## Results

Eleven clinicians and 15 caregivers took part in the study. Almost all caregivers identified as women (93%) and as non-Hispanic White (93%). Almost half (47%) were between the ages of 30 and 49 years old, and all but one were adoptive parents (93%; Table 1).

**Table 1.** Sociodemographic characteristics of caregivers and children

| Characteristics | % (N) |
| --- | --- |
| <u>Gender</u> |  |
| Man | 7% (1) |
| Woman | 93% (14) |
| Transgender/other | 0% (0) |
| <u>Race/Ethnicity</u> |  |
| Non-Hispanic White | 93% (14) |
| Non-Hispanic Black/African-American | 7% (1) |
| Hispanic/Latinx, any race | 0% (0) |
| <u>Age</u> |  |
| 30-49 Years Old | 47% (7) |
| 50-59 Years Old | 33% (5) |
| +60 Years Old | 20% (3) |
| <u>Relationship to Child</u> |  |
| Adoptive Parent | 93% (14) |
| Foster Parent | 0% (0) |
| Other | 7% (1) |
| **Characteristics of child diagnosed with FASD** | **N (%)** |
| <u>Current Age of Child</u> |  |
| <5 Years Old | 29% (4) |
| 5-12 Years Old | 50% (7) |
| 13-17 Years Old | 21% (3) |
| <u>Age at time of FASD Diagnosis</u> |  |
| Less Than 5 Years Old | 50% (7) |
| 5-12 Years Old | 50% (7) |
| 13-17 Years Old | 0% (0) |
| <u>Child's Insurance</u> |  |
| Medicaid | 86% (12) |
| Private | 7% (1) |
| Medicaid & Private | 7% (1) |
| Other | 0% (0) |

Caregivers each reported on one child under their care who had been diagnosed with FASD. Half of these children had been diagnosed with FASD when they were younger than five years old (50%). Most children were covered by Medicaid (86%).

Almost all clinicians were women (91%) and most were non-Hispanic White (64%). Nearly half were psychologists (45.5%). Almost half had treated 11-99 patients suspected of having FASD in the 12 months prior to being screened (45.5%; Table 2).

**Table 2.** Sociodemographic characteristics of the clinician sample

| Table 2. Sociodemographic characteristics of the clinician sample |  |
| --- | --- |
| Characteristics | % (N) |
| <u>Gender</u> |  |
| Man | 9% (1) |
| Woman | 91% (10) |
| Transgender/Other | 0% (0) |
| <u>Race/Ethnicity</u> |  |
| Non-Hispanic White | 64% (7) |
| Hispanic/Latino | 27% (3) |
| Asian-American | 9% (1) |
| <u>Age</u> |  |
| 30-54 | 45.5% (5) |
| 55-79 | 45.5% (5) |
| 80+ | 9% (1) |
| <u>Profession</u> |  |
| Psychologist | 46% (5) |
| Psychiatrist | 18% (2) |
| Consultant | 9% (1) |
| Education Specialist | 9% (1) |
| Neurologist | 9% (1) |
| Pediatrician | 9% (1) |
| <u>Years Practicing in Profession</u> |  |
| 1-9 | 27.3% (3) |
| 11-19 | 9% (1) |
| 20-29 | 18.2% (2) |
| +30 | 45.5% (5) |
| <u>Number of FASD Patients Seen in Past 12 Months</u> |  |
| 1-10 | 36.4% (4) |
| 11-99 | 45.5% (5) |
| 100+ | 18.1% (2) |

The qualitative analysis identified three themes: (1) precipitating concerns and screening; (2) barriers to FASD evaluation and diagnosis; and (3) barriers and facilitators to linking to FASD-related interventions and resources. Several themes had salient sub-themes (see Figure 1).

**Figure 1.**
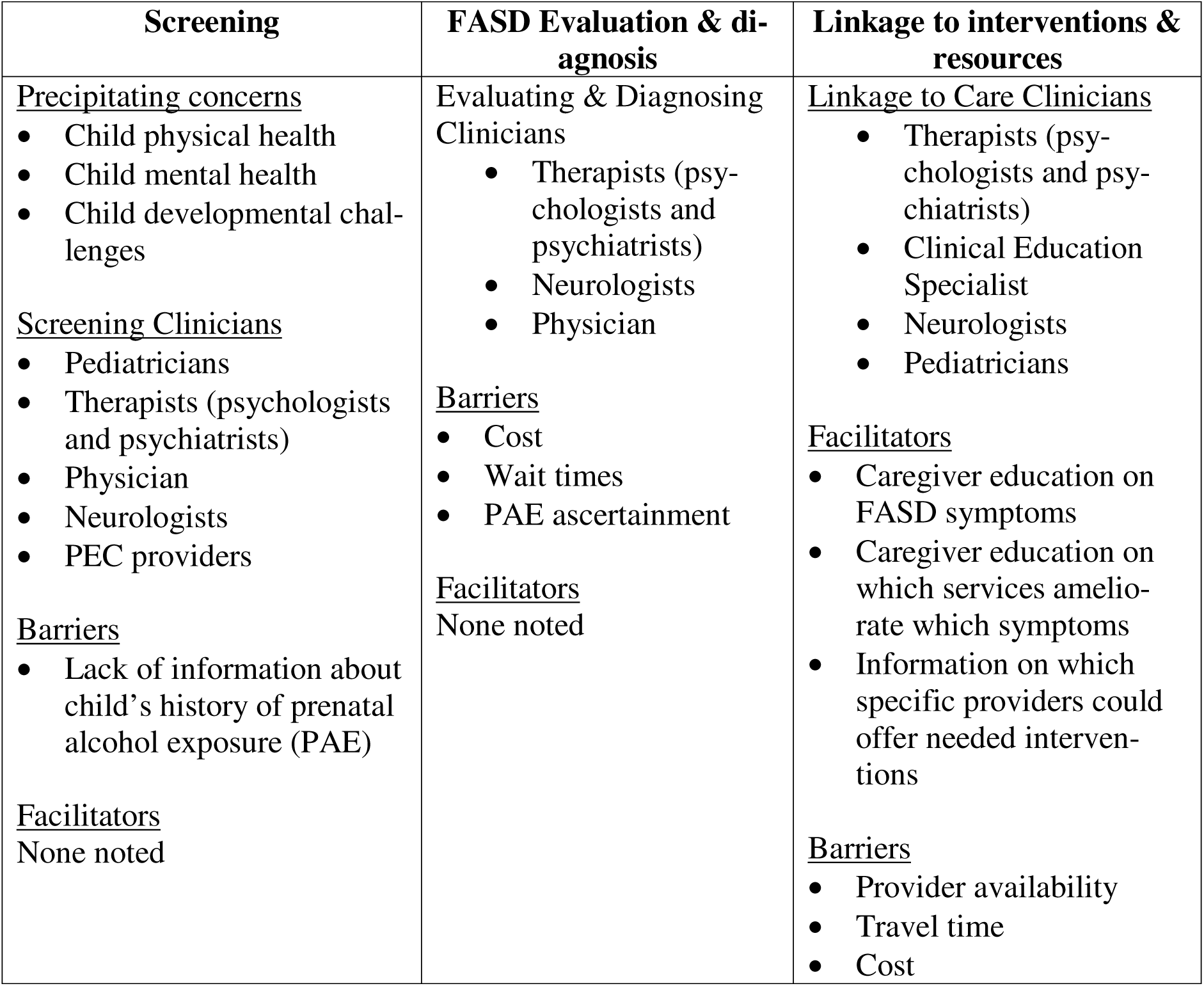
Themes and subthemes identified in the analysis

### Precipitating Concerns and Screening

Caregivers typically initiated screening out of concern for the child’s physical or mental health or developmental challenges (e.g., behavioral issues). Sometimes they were unaware that prenatal alcohol exposure [PAE] might have caused these issues. In most cases, screenings were conducted by pediatricians, therapists, including psychologists and psychiatrists, and neurologists; children needing further evaluation were then referred to the PEC.

The primary barrier to screenings was absence of information about PAE. This barrier is discussed in detail in the next section.

### Barriers to FASD Evaluation and Diagnosis

Both caregivers and clinicians reported that obtaining an evaluation and subsequent diagnosis could be exceptionally challenging. The most commonly discussed barriers were (1) the cost of the evaluation; (2) wait times; and (3) PAE documentation. We discuss each of these barriers in the order in which caregivers experienced them during the FASD diagnostic journey. The analysis identified no facilitators to securing an evaluation or diagnosis.

#### Evaluation Cost

FASD evaluations have multiple components, including but not limited to physical, neurological, psychological, and cognitive assessments; providers with specialized training must conduct these assessments. As a result, clinicians and caregivers reported that FASD evaluations are costly and that this cost was a major barrier. Caregivers and clinicians reported that Medicaid and other insurance plans often covered only a portion of the evaluation or none at all. Paying out-of-pocket was out of the question for many families: collectively, these evaluations cost approximately $2000. Costs increased if more tests were needed to finalize a diagnosis (e.g., genetic testing). Even if Medicaid covered the evaluation, many providers refused to accept it. A caregiver said:

> *“nobody wants to take [Medicaid], so…you just get kicked around from place to place and it’s just so unfair because what are you going to do? You’re gonna pay $2,000, $1,800 [out of pocket]…that was the price that I was quoted for…having…an assessment done”*

A clinician explained why: “it costs about $2000 for an evaluation and Medicaid will pay $148.” The PEC was able to accept patients who were on Medicaid or were uninsured because the state legislature funded them to work with this population. This funding was, however, limited and the PEC thus had to restrict the number of children evaluated. As one clinician summarized, “the biggest barrier [to diagnosis]… is the insurance companies.”

#### Wait times

Clinicians and caregivers reported that wait times for FASD evaluation ranged from six weeks to two years, with yearlong delays being routine. Delayed evaluations had roots in both clinician constraints and caregivers’ lives. The demand for evaluation far outstripped local capacity; providers with the skills needed to conduct FASD evaluations were rare in this Georgia metropolitan area. Clinicians explained that the few clinics that were equipped to undertake these complex evaluations were understaffed: because of inadequate insurance reimbursement for evaluations, clinics could not afford to hire providers with needed expertise.

Delays also originated in families. Some caregivers were raising several medically complex children and the FASD evaluation was not the most urgent concern. As one caregiver said,

> *“We had to reschedule one of the evaluation appointments because another kid was in the hospital… what are you gonna do?… it took …longer[to reschedule] than I wanted…”*

Wait times were compounded when multiple appointments with different providers were needed to finalize the evaluation process.

Diagnostic delays caused anxiety about child wellbeing. As one caregiver reported:

> *“if [FASD] is going on, we need to have it diagnosed because there were … milestones that weren’t being met and… if we get this diagnosed early, the better chances of us being able to work with it through the school system and, like, just [get] resources in place that are needed… If you don’t have a diagnosis you can’t go anywhere…”*

#### Documented Confirmation of PAE

Caregivers and clinicians reported that accurately ascertaining PAE was an essential component of the FASD diagnostic process but was exceptionally challenging. Clinicians explained that acceptable PAE confirmation included (1) verbal disclosure by the birthmother to the clinician; (2) written disclosure by either birthparent; (3) eyewitness report; or (4) medical/birth record or legal/social service documentation. As one clinician noted,

> *“You can’t say ‘someone told me’… [The PAE report] can’t be secondhand. It’s something someone had to observe [if the birthparent doesn’t report it directly]…”*

Stigma, absent birthparents, and poor record keeping by relevant agencies often rendered it impossible to confirm PAE. Participants observed that alcohol consumption during pregnancy is heavily stigmatized and could precipitate state child protective service engagement. As a result, birthparents could be exceptionally and understandably reluctant to disclose. A clinician said,

> *“…. a woman who’s a biological mother and who’s been sent here by the courts, she may or may not…give us an accurate statement about her [alcohol] use…that’s going to depend where her head is. If she’s in recovery, she may tell us exactly what happened …..people are under a variety of legal and other constraints and they’ll tell you what seems to be the best [under these constraints] at the time.”*

When neither birthparent was present at the evaluation, other caregivers who were present often had not personally witnessed the birthmother consume alcohol, and so could not provide an eyewitness account. Moreover, caregivers could be reluctant to disclose PAE if the birthparents were in the midst of a child protective service investigation or other custody actions.

Poor record keeping and suboptimal access to records formed a third barrier to PAE confirmation. Clinicians reported that obstetric and birth records often did not contain information about birthmother alcohol use. Obstetricians often did not query alcohol consumption, and even when pregnant people were asked about their use and confirmed it, the information was not recorded. Even when information on PAE was queried and recorded, the resulting records might be impossible to obtain if the encounter had occurred outside of the university healthcare system, particularly if that external system used a different electronic health record platform.

Multiple caregivers expressed frustration with the insistence on PAE as a diagnostic criterion for FASD. They believed that it unnecessarily prevented children with FASD from securing needed services. As one caregiver said,

> *“… [The FASD evaluation clinic] said… our oldest daughter would have [been diagnosed with FASD] if there was proof that mom drank… she checked all the boxes except that one. Mom wouldn’t admit it…[our daughter] has some things going on, but she didn’t really check enough boxes for anything.”*

#### Barriers and Facilitators to Linking to Interventions

The most commonly mentioned FASD-related services needed were occupational therapy, speech therapy, and academic support. While securing a diagnosis was a necessary precondition for accessing these and other needed services, it was often not sufficient. Participants identified facilitators and barriers to connecting with intervention services. Facilitators were caregiver education surrounding FASD and receipt of intervention recommendations. Unfortunately, caregivers and clinicians noted that recommended services were often unavailable, far away, or too costly.

### Facilitators: Caregiver Education and Recommended Services

When their child received the FASD diagnosis, caregivers often lacked knowledge about FASD symptoms and how they could be treated, about which services might ameliorate which symptoms, and about how to access these services. Clinician guidance was thus essential. During the visit in which the diagnosis was disclosed, clinicians informed caregivers about services their children needed to address their specific constellation of symptoms, and shared contact information of specific providers of those services. One clinician said,

> *“Providing explicit names, telephone numbers, and explicitly telling the caregivers why this recommendation has been made and what the anticipated outcome may be [is essential]…and [explaining] why it’s beneficial for the child. But the biggest thing is if we… [give] them …the website [of the service]…’here’s who you need to contact.’ [If we do this,] they’re more apt to get the support and services”*

As one caregiver said,

> *”…the team …helped us look for therapists when the time came…was able to [share] more information on the therapy place [and] whether that might be a good fit …”*

Caregivers often called out Educational Advocates as essential partners in connecting children to needed academic services. This Advocate identified specific services that the child needed, based on their evaluation results, and then advocated to ensure the child received them through the school whenever possible. In this quote, a caregiver describes how the PEC’s Educational Advocate helped the school understand the child’s academic needs: The Advocate

> *“…[sat in] on one of our IEP [meetings and]…she was helpful in helping them understand how much extra help he needed and…making suggestions to the school [on how to provide this help].”*

#### Barriers: Non-existent services, long travel times, and cost

Unfortunately, families routinely could not engage with the recommended services – academic or otherwise – because the services did not exist at all, were too far away, or were prohibitively expensive. The most commonly reported barrier was simply that no provider of the needed services practiced within a reasonable travel distance of the child’s home. Psychologists/counselors and academic supports were particularly hard to find. As one caregiver stated:

> *“There’s really not enough services for these kids…we know we can get some OT [i.e., occupational therapy], we can get some speech, we can get an IEP (Individualized Education Plan). We don’t get much past that.”*

The availability of school-based services varied by locality. One clinician noted,

> *“I once had a kid who couldn’t get an occupational therapist… in [X] county and the school system had, for two years, [tried] to get an occupational therapist in their school system.. [but hadn’t been able to].”*

Several participants reported that the service environment was highly dynamic, because budget cuts often closed services down. This made referrals challenging.

> *“Some of the services that originally were in place and that you refer families to, the funding stops.”*

Travel times to distant providers could be prohibitive, given obstacles experienced by families. As one clinician commented,

> *“We [might] refer them to a therapist… but they would never make it to that appointment [because of] transportation issues…they couldn’t get time off from work [to travel]… they wouldn’t get [the services] they needed.”*

Long travel distances were particularly common for rural families, with some traveling an hour or more to access weekly or biweekly services because needed services were unavailable locally. For example, one clinician explained that,

> *“The only thing the [rural] family could do was drive to <u>[city]</u> which was like an hour away from where they lived and it was just a huge hassle [to make that trip weekly].”*

As with evaluations, cost was a persistent issue because providers refused Medicaid.

Collectively, these barriers meant that families and children routinely did not get needed services. As one clinician noted,

> *“I…go over the previous recommendations [at each follow-up appointment] to see… what gains have been made, and nine [times] out of ten those recommendations are not followed through… it could be many reasons. It’s not that they don’t want to do it. It’s just ‘oh we called and they no longer take our insurance’ or ‘that’s too far’.*

## Discussion

Amidst increasing PAE in the US, this analyses revealed several significant barriers and facilitators within this diagnostic and intervention journey. Cost, long wait times, and poor PAE documentation impeded FASD diagnosis. Diagnostic delays ranged from six weeks to two years and raised concerns about missed opportunities to engage with transformative early interventions. Clinician education of caregivers supported linkage to FASD-related interventions, but this education often could not overcome the absence of local providers, cost, and travel times.

Cost was a recurring impediment to both FASD diagnoses and linkage to interventions. Because they require physical, neurological, psychological, and cognitive assessments, evaluations cost a minimum of $2000. Caregivers and clinicians reported that Medicaid – the most commonly reported insurer of children in this sample and of children with FASD in the US as a whole – did not cover some or any of these assessments. In the US, unfortunately, federal funding for FASD prevention and intervention has lagged behind increasing need. For example, the FASD Prevention Act of 1998 allocated $27 million to FASD-related prevention and intervention, and this figure declined to $12 million in 2021. (FASD United ND) Advocacy by people living with FASD, their caregivers, and providers for the Federal FASD Respect Act holds promise: this bill includes provisions to fund states and Tribal Systems to provide FASD services across the lifespan.(FASD United ND)

Provider availability curtailed both FASD evaluation and diagnosis and engagement with interventions. Advocates have long warned of this care crisis,(FASD United ND) and recent systematic review supports these experiences: awareness and knowledge of FASD itself is poor among healthcare providers (e.g., 4% of midwives are aware of PAE and its consequences), as is experience evaluating and treating children for FASD.(O’Connor et al. 2022) Awareness and knowledge of FASD is low even among psychologists, who often treat people living with FASD and are well-positioned to diagnose this disorder.(O’Connor et al. 2022) Low awareness and knowledge among providers, coupled with stigmatizing attitudes toward PAE and its consequences,(Choate and Badry 2019) limited Medicaid reimbursement, significantly impede the FASD evaluation, diagnostic, and linkage to intervention journey for children living with FASD and their families. Addressing evaluation, diagnostic, and linkage to care capacity in US medical systems is a priority for national advocacy groups, and collectively form a stipulation in the FASD Respect Act.(FASD United ND)

### Limitations and Suggestions for future research

This study’s limitations inform next steps for future research. Our sample was homogeneous by virtue of its affiliation with a single, albeit large, hospital system with a specialty neuro-developmental clinic serving one Georgian metropolitan area. Heterogeneities exist in provider availability across states and hospital systems; in organizational policies guiding PAE screening and recording; in state and facility responses to positive PAE screens; and across states in the services that Medicaid and other insurers must cover for children. Sister qualitative studies in other US settings might offer contrasting results, and differences across settings could identify key policy and organizational interventions to improve the FASD diagnostic journey.

Our sample of caregivers was homogenous: most were adoptive parents and were White. These characteristics are not completely representative of those receiving care at PEC. Willingness to query, report, and record PAE varies across racial/ethnic groups because of racialized enforcement of laws governing substance use in pregnancy among other issues.(Cooper et al. 2025)

Moreover, adoptive parents could not speak to experiences of disclosing PAE. All had received diagnostic services from PEC, and thus we could not explore variations in diagnostic experiences across facilities (including non-specialty providers) or explore the experiences of caregivers who suspected their children had FASD but had never been able to secure an evaluation or diagnosis. Future studies should be conducted with foster and biological parents, with more racially and ethnically diverse caregiver populations, and with caregivers seeking care at non-specialty providers to explore how these features shape the FASD diagnostic journey.

## Conclusion

Amidst rising PAE, this sample of clinicians and caregivers reported that cost, provider availability, long wait times, and PAE documentation impeded FASD evaluation, diagnosis, and linkage to services. Past research supports these findings, and identifies structural drivers of identified barriers, including low Medicaid reimbursement rates, declining federal funding for FASD-specific services, and poor awareness and knowledge of FASD among relevant healthcare providers. The persistence and success of ongoing advocacy efforts among people living with FASD, their caregivers, and providers provides essential pathways toward addressing these barriers.

## Data Availability

All data produced in the present study are available upon reasonable request to the authors

## Acknowledgements

We are grateful to the individuals who took part in this project, and to Drs. Nicholas Deputy, Mary-Kate Weber, and Ali Ruprecht, and to Ms. Micah Wilkins and Caitlin Green for their leadership of the multisite qualitative project. This project was supported by the Centers for Disease Control and Prevention (CDC) of the U.S. Department of Health and Human Services (HHS) as part of a financial assistance award totaling $1,163,850 with 100% funded by CDC/HHS. The contents are those of the author(s) and do not necessarily represent the official views of, nor an endorsement, by the U.S. Government.

